# Stranded abroad during the COVID-19 pandemic: examining the psychological and financial impact of border restrictions

**DOI:** 10.1101/2021.12.08.21267218

**Authors:** Pippa McDermid, Adam Craig, Meru Sheel, Katrina Blazek, Siobhan Talty, Holly Seale

## Abstract

**Objective:** With the easing of COVID-19-related international travel restrictions in late 2021 it is time to consider the direct and indirect social, emotional, and financial impacts that these border closures have had. The study aims to evaluate the psychological and financial distress reported by people stranded abroad due to international travel restrictions introduced in response to the COVID-19 pandemic.

**Methods:** Between July and September 2021, we implemented a cross-sectional online survey targeting individuals stranded abroad due to international travel restrictions. The survey collected data about COVID-19 travel restriction-related travel impacts; personal stress, anxiety, and depression (using the validated DASS-21tool); as well as impacts on housing and financial security; and demographic data.

**Findings:** We had 1054 participants complete the survey; most were trying to return to the Oceania region (75.4%), with 45% stranded in Europe. Overall, 64.2% reported financial distress while stranded abroad. 64.4% (x̄ =9.43, SD=5.81) reported either a moderate or severe (based on the DASS-21 classification) level of depression, 41.7% for anxiety (x̄ =5.46, SD=4.74), and 58.1% for stress (x̄ =10.64, SD=5.26). Statistically significant factors associated with moderate to extremely severe depression, anxiety, and stress were financial stress, an employment change, being <30yrs, having a high perceived risk of contracting COVID-19 abroad and being stranded for >2 months.

**Conclusion:** The study is among the first to explore the psychological and financial distress-related impacts associated with being stranded abroad due to COVID-19 travel restrictions. It highlights a range of unintended consequences that arise from pandemic-related travel restriction, identifies the health and social needs for a particularly vulnerable population, and provides clues as to the types of support that may be adopted to best support them. This research will assist policymakers in identifying support packages for people stranded abroad due to global disaster.

## BACKGROUND

In response to the COVID-19 pandemic, most countries around the world have implemented some level of international travel restriction or complete border closures (1-3). As of February 2020, many countries had commenced repatriation of their citizens stranded abroad. By the end of 2020, some countries like Japan and Spain claimed to have completely repatriated every citizen that wanted to return. However, reports continue to suggest stranded travellers are still trying to get to their country of residence (referred to as ‘home’) 20 months into the COVID-19 pandemic despite many countries reopening borders (4, 5).

Factors impacting on people’s ability to return to their home have included countries placing limits on the number of passengers that can enter the country, caps on the hotel quarantine capacities, the cost of travel and hotel quarantine, and in some cases having restrictions on flights from certain high-risk countries. Our previous study (pre-print available) suggested that the support available to those who were stranded abroad was limited, and in some cases difficult to access and comprehend (6). Support provided by countries has varied from repatriation flights, emergency accommodation, mental health and medical assistance, emergency call lines and financial assistance. However, of the countries that were reviewed, we were unable to identify any one country providing all the different support types listed. Public commentary through news and social media has hinted at the level of psychosocial impact on citizens stranded abroad. These articles suggest that many of these travellers felt abandoned by their governments, had little financial support, and for some, experienced depression and homelessness (7-9). While the findings from many COVID-19 studies have reported high levels of psychological distress in nearly all populations, the focus of these studies has been on domestic populations, like healthcare and frontline workers, students and those in lockdown and quarantine (9-18). One study, with similar aims to the present study, found 63% of Saudi citizens living abroad as students during COVID-19 experienced ‘psychiatric’ distress symptoms (19).

There is currently limited understanding of the level of psychological distress that has been experienced by those stranded abroad wanting to return. This study examined the impacts of travel restrictions on people stranded abroad, who were unable to return to their country of citizenship/residence during the COVID-19 pandemic. We aimed to 1) measure the prevalence of psychological impact associated with being stranded overseas due to covid-related travel restrictions and 2) identify demographic and circumstantial factors associated with severe psychological impact.

## METHODS

### Population and procedures

An online survey was created and administered anonymously using the *Qualtrics* (20) survey platform, with respondents recruited through a variety of social media channels. Respondents were those people who were either still stranded away from their country of residence/home or had been stranded at some point since the commencement of the COVID-19 pandemic. There were no limitations placed on the country of residence, nor the length of time the person had been stranded for. To meet the inclusion criteria, respondents had to self-identify as having attempted to return to their country of residence but have had their travel plans changed. The survey was open between 20 July 2021 and 24 September 2021. Participants unique IP addresses prevented duplicate entries. Ethical approval for this study was granted by the UNSW Human Research Ethics Committee (#210418). All participants indicated their consent to participate.

### Survey instruments and measures

Demographics: included gender, age, level of education, ethnicity, employment status, history of chronic illness, and living status. Ethnicity was classified based on the nine broad groups according to the Australian Standard Classification of Cultural and Ethnic Groups (21).

Travel experiences: Respondents were asked where they were stranded abroad and where they intended to return to, and this data was recoded into geographic groups based on the World Health Organisation regions (22). Questions focused on their current situation (whether they had returned, were still stranded abroad awaiting return or still abroad but had decided to stay), flight cancellations/delays, length of time waiting to return and their experiences with travel.

Mental wellbeing: The depression, anxiety, and stress scale (DASS-21) was used in this study (23). The DASS-21 is a validated self-report tool, previously used in COVID-19 research studies (13, 24), containing 21 items assessing scores of depression, anxiety, and stress symptoms (7 items each). Respondents were asked to reflect on when they were stranded abroad (for participants who had already returned) or reflecting on the last two weeks and rate each statement on a 5-point Likert scale from 0 (unsure/do not recall; did not apply to me at all) to 3 (applied to me very much, or most of the time). Scores in each sub section are then multiplied by 2 to give a final score categorising the depression, anxiety, and stress into normal, mild, moderate, severe, or extremely severe. Higher scores reflect increased emotional and psychological distress.

As we suspected some potential participants having returned home already and could be potentially reflecting further than 2 weeks, we included an additional option of “unsure/do not recall” to allow for removal of recall bias during the analyses phase. Participants were asked whether they had access to crisis support or mental health services while abroad and if ‘yes’, whether they had used this service or support. Finally, respondents reported their perceived risk of contracting COVID-19 both in the country where they are/were stranded and the country where they had or were waiting to return to (Scale of 1-10; 1 being no risk and 10 being high risk).

Financial wellbeing: Respondents were initially asked whether they felt financial stress while stranded abroad (yes/no), then if ‘yes’ were asked how they addressed the financial stress (receiving financial support from family, government loans, bank credit, or social services). Questions on employment situation and changes while abroad were asked along with a question on whether the participant experienced homelessness while abroad. Homelessness was defined as a period where participants did not have somewhere to stay/live.

### Statistical analyses

Descriptive analysis involved the calculation of means, standard deviation, confidence interval and standard errors. Chi-square test of independence were first used to compare categorical variables. Independent variables that showed a significant association with DASS severity scores at a *p*<.2 level was included in the model as predictor variables. The DASS scores were dichotomised to reflect either no/mild symptom severity or moderate to extremely severe. Multivariable logistic regression was then performed to analyse the effect of age, perceived risk of contracting COVID-19 while stranded, financial stress, time stranded, employment change and homelessness on predicting moderate to extremely DASS. No multicollinearity amongst variables was identified. P values of <.05 was considered statistically significant. All data analyses were conducted using SPSS (25).

## RESULTS

### Participant characteristics

A total of 1054 participants completed the full survey, while a further 296 respondents completed over 75% of the questions and were included. Demographic information is provided in Table 1. The mean age was 41.09±13.08, with 69.5% being female, 43.8% of north-west European ethnicity, while most had a tertiary education (90.7%) and were stranded in the European Region (EUR) and were trying to return to the Western Pacific Region (WPR).

**Table 1.**
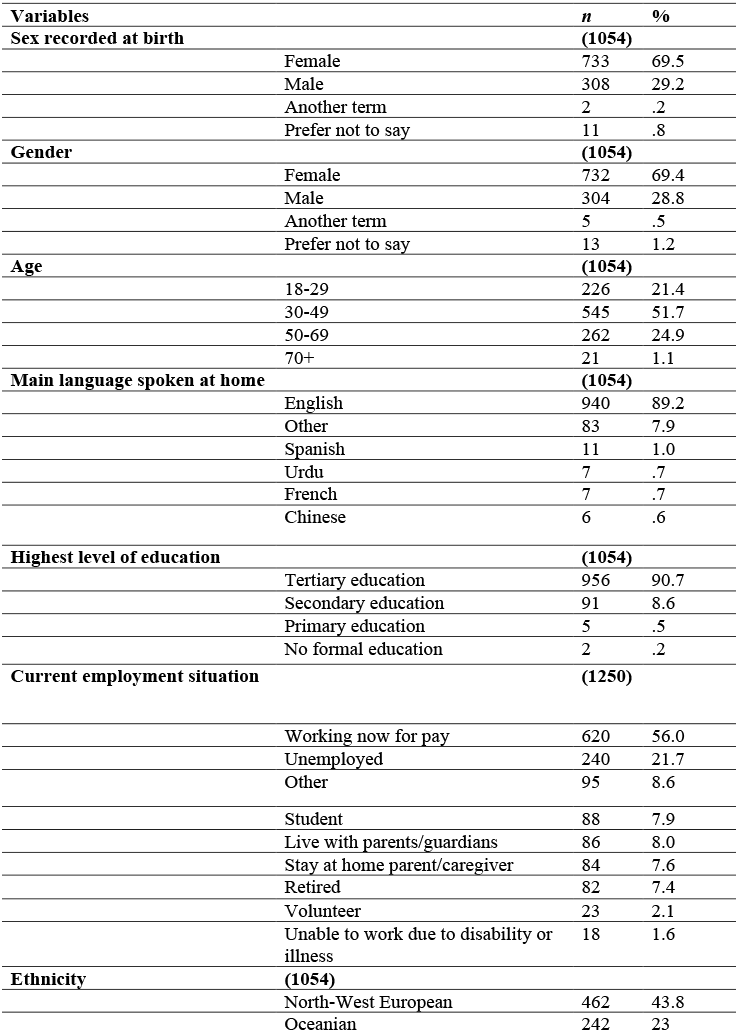
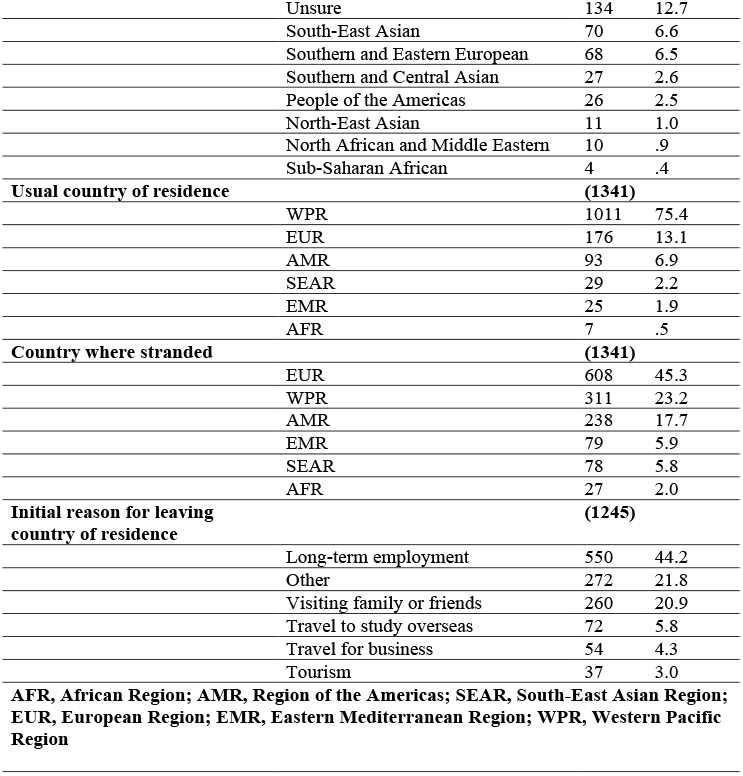
Characteristics of participants stranded during COVID-19.

| **Variables** | ***n*** | **%** |
| --- | --- | --- |
| **Sex recorded at birth** | **(1054)** |  |
| Female | 733 | 69.5 |
| Male | 308 | 29.2 |
| Another term | 2 | .2 |
| Prefer not to say | 11 | .8 |
| **Gender** | **(1054)** |  |
| Female | 732 | 69.4 |
| Male | 304 | 28.8 |
| Another term | 5 | .5 |
| Prefer not to say | 13 | 1.2 |
| **Age** | **(1054)** |  |
| 18-29 | 226 | 21.4 |
| 30-49 | 545 | 51.7 |
| 50-69 | 262 | 24.9 |
| 70+ | 21 | 1.1 |
| **Main language spoken at home** | **(1054)** |  |
| English | 940 | 89.2 |
| Other | 83 | 7.9 |
| Spanish | 11 | 1.0 |
| Urdu | 7 | .7 |
| French | 7 | .7 |
| Chinese | 6 | .6 |
| **Highest level of education** | **(1054)** |  |
| Tertiary education | 956 | 90.7 |
| Secondary education | 91 | 8.6 |
| Primary education | 5 | .5 |
| No formal education | 2 | .2 |
| **Current employment situation** | **(1250)** |  |
| Working now for pay | 620 | 56.0 |
| Unemployed | 240 | 21.7 |
| Other | 95 | 8.6 |
| Student | 88 | 7.9 |
| Live with parents/guardians | 86 | 8.0 |
| Stay at home parent/caregiver | 84 | 7.6 |
| Retired | 82 | 7.4 |
| Volunteer | 23 | 2.1 |
| Unable to work due to disability or illness | 18 | 1.6 |
| **Ethnicity** | **(1054)** |  |
| North-West European | 462 | 43.8 |
| Oceanian | 242 | 23 |

|  |  |  |  |
| --- | --- | --- | --- |
|  | Unsure | 134 | 12.7 |
|  | South-East Asian | 70 | 6.6 |
|  | Southern and Eastern European | 68 | 6.5 |
|  | Southern and Central Asian | 27 | 2.6 |
|  | People of the Americas | 26 | 2.5 |
|  | North-East Asian | 11 | 1.0 |
|  | North African and Middle Eastern | 10 | .9 |
|  | Sub-Saharan African | 4 | .4 |
| **Usual country of residence** |  | **(1341)** |  |
|  | WPR | 1011 | 75.4 |
|  | EUR | 176 | 13.1 |
|  | AMR | 93 | 6.9 |
|  | SEAR | 29 | 2.2 |
|  | EMR | 25 | 1.9 |
|  | AFR | 7 | .5 |
| **Country where stranded** |  | **(1341)** |  |
|  | EUR | 608 | 45.3 |
|  | WPR | 311 | 23.2 |
|  | AMR | 238 | 17.7 |
|  | EMR | 79 | 5.9 |
|  | SEAR | 78 | 5.8 |
|  | AFR | 27 | 2.0 |
| **Initial reason for leaving country of residence** |  | **(1245)** |  |
|  | Long-term employment | 550 | 44.2 |
|  | Other | 272 | 21.8 |
|  | Visiting family or friends | 260 | 20.9 |
|  | Travel to study overseas | 72 | 5.8 |
|  | Travel for business | 54 | 4.3 |
|  | Tourism | 37 | 3.0 |
| **AFR, African Region; AMR, Region of the Americas; SEAR, South-East Asian Region; EUR, European Region; EMR, Eastern Mediterranean Region; WPR, Western Pacific Region** |  |  |  |

Almost one in four participants (*n* = 303/1214) reported a historical or current COVID-19 infection, and of those the majority rated a ‘mild’ symptom severity (85.1%). Participants’ overall level of perceived risk of contracting COVID-19 while abroad was moderate to high (*n* =806, M = 6.64, *SD*=2.85, range 1-10, where 1 = no risk, and 10=high risk), with 25.4% rating the perceived risk while abroad at high risk (10). Comparatively, the overall level of perceived risk of contracting COVID-19 in the country where participants had returned to was low to moderate (*n* =605, M = 5.10, *SD*=2.72, range 1-10, where 1 = no risk, and 10=high risk), with 21.4% rating the perceived risk at ‘home’ as low risk.

### Travel experiences

Initially 44% of respondents had left their country of residence to take up long-term employment, with over 60% stranded abroad for more than 5 months (63.6%), and just over a quarter either having had booked a flight or awaiting flight availability (26.6%). See table 3 for a full breakdown of participants experiences and current situation while stranded abroad.

**Table 2.** Travel experiences of participants stranded abroad during COVID-19.

| **Variables** | ***n*** | **%** |
| --- | --- | --- |
| **Time stranded abroad** | **(1341)** |  |
| <1 month | 89 | 6.6 |
| 1-2 months | 91 | 6.8 |
| 2-3 months | 128 | 9.5 |
| 3-5 months | 98 | 7.3 |
| >5 months | 853 | 63.6 |
| No wait | 82 | 6.1 |
| **Number of flight cancellation/changes** | **(1245)** |  |
| Not applicable | 319 | 25.6 |
| 0 changes | 152 | 12.2 |
| 1 cancellation or delay | 245 | 19.7 |
| 2 cancellation or delay | 196 | 15.7 |
| 3 cancellation or delay | 147 | 11.8 |
| 4 cancellation or delay | 65 | 5.2 |
| 5 cancellation or delay | 31 | 2.5 |
| >5 cancellations or delays | 90 | 7.2 |
| **Experiences while trying to return** | **(1245)** |  |
| Limit on the number of people who could enter the country | 740 | 59.4 |
| Inability to book a flight | 671 | 53.9 |
| Flight cancellation | 660 | 15.3 |
| COVID-19 testing requirements prior to flying | 281 | 22.6 |
| Other | 229 | 18.4 |
| Separation from family/companion | **172** | 13.8 |
| Visa issues | 147 | 11.8 |
| Delays during transit | 44 | 3.5 |
| **Current situation** | **(1245)** |  |
| Booked a flight/waiting for flight availability | 357 | 26.6 |
| Unable to return but have decided to stay | 352 | 26.2 |
| Returned to country of residence after delays | 323 | 24.1 |
| Other | 295 | 22 |
| Returned to country of residence after being stuck in transit | 14 | 1 |

**Table 3.** Financial and employment characteristics of participants stranded abroad during COVID-19.

| **Variables** | ***n*** | **%** |
| --- | --- | --- |
| **Addressing financial stress** | **(763)** |  |
| Received financial support from family or friends | 354 | 46.4 |
| Borrowed money from a bank | 58 | 7.6 |
| Received a government loan to cover your living costs | 14 | 1.8 |
| Received a government loan to cover the cost of a flight home | 8 | 1 |
| Accessed emergency financial support from the organisation or services in the country you were stuck | 43 | 5.6 |
| Received financial support from an insurance company | 6 | .8 |
| Received financial support from your employer | 39 | 5.1 |
| Applied but did not receive a government loan | 39 | 5.1 |
| Personal savings | 47 | 6.2 |
| Early withdrawal of superannuation | 27 | 3.5 |
| Other | 128 | 16.8 |
| **Current employment situation** | **(1250)** |  |
| Working now for pay | 620 | 56.0 |
| Unemployed | 240 | 21.7 |
| Retired | 82 | 7.4 |
| Student | 88 | 7.9 |
| Unable to work due to disability or illness | 18 | 1.6 |
| Volunteer | 23 | 2.1 |
| Stay at home parent/caregiver | 84 | 7.6 |
| Other | 95 | 8.6 |
| **Employment change while abroad** | **(717)** |  |
| Worked remotely | 123 | 17.2 |
| Lost job | 119 | 16.6 |
| Stood down, not working for pay, but not fired | 45 | 6.3 |
| Pay cut | 32 | 4.5 |
| Reduction of hours | 63 | 8.8 |
| Not working but receiving government assistance | 27 | 3.8 |
| Back in paid work | 31 | 4.3 |
| Resigned | 77 | 10.7 |
| Contract not renewed | 69 | 9.6 |
| Increase in hours | 18 | 2.5 |
| Other | 113 | 15.8 |

### Mental wellbeing

Figure 1 presents the respondents (*n*=1133) self-reported depression, anxiety, stress symptom severity scores based on the DASS-21 tool. Of the respondents, 64.4% scored moderate to extremely severe depression symptoms 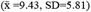, 41.7% scoring moderate to extremely severe anxiety symptoms 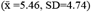, and 58.1% scoring between moderate to extremely severe stress symptoms 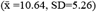.

**Figure 1.**
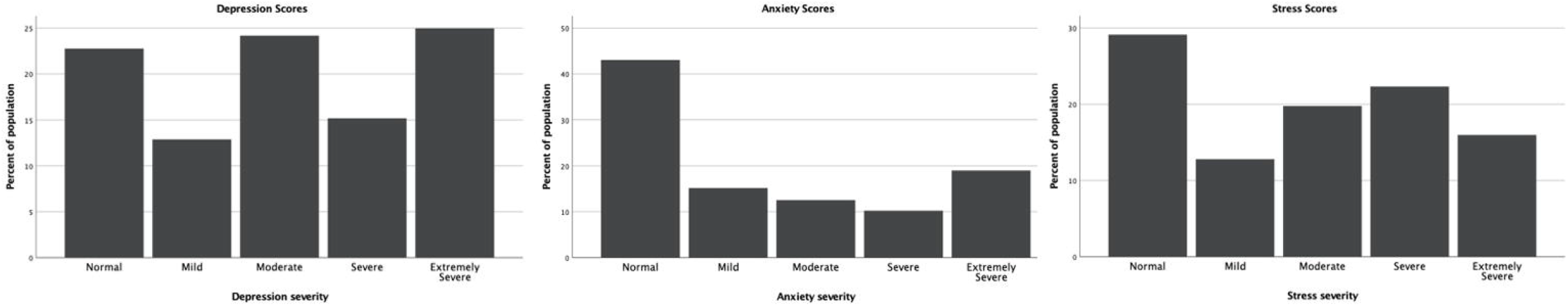
Depression, anxiety, and stress categories of citizens stranded abroad during COVID-19 (% of sample)

Many reported no access to crisis support or mental health services while abroad (63.5%) (*n* = 719/1133), and of those that did have access, only 37.9% (*n* = 272/719) utilised the services. A total of 12% (*n* = 133/1112) experienced a period of homelessness while stranded abroad. Of those that were willing to share their experiences (*n* = 94), commonly noted situations included living in temporary accommodation (32%), sleeping on the couch or in a spare bedroom at a friend/family members place (32%), and staying in emergency accommodation (including homeless shelters) (17%). Less common experiences were those that lived on the street, trains, at the airport, in cars, in tents (<20%). Two respondents disclosed having experienced a sexual assault while homeless shelter.

### Financial wellbeing

Financial distress was reported in 64.2% (n=723/1127), and 45% (n=433/1127) reported a change in employment. A breakdown of ways in which participants sought to address financial distress along with employment changes and current employment situation are reported in Table 3.

### Factors associated with and predictors of depression, anxiety, and stress

Chi square analyses revealed significant associations between participants DASS categories and their age, time stranded abroad, financial stress, homelessness, employment change, and their perceived risk of contracting COVID-19. No associations were found between DASS severity categories and having access to crisis support or mental health services (see Supplementary file 1). For depression, logistic regression identified financial stress, employment change, and a high perceived risk of contracting COVID-19 as predictors of moderate to extremely severe depression. Overall, the model correctly discriminated 67.7% of cases and Nagelkerke R^2^ indicated a 14% variation of depression explained by the model. For anxiety, logistic regression identified financial stress, employment change, and a high perceived risk of contracting COVID-19, as predictors of moderate to extremely severe anxiety. Overall, the model correctly discriminated 64.5% of cases and Nagelkerke R^2^ indicated a 13% variation of anxiety explained by the model. Finally for stress, logistic regression identified financial stress, employment change, and a high perceived risk of contracting COVID-19, as predictors of moderate to extremely severe stress. Overall, the model showed goodness of fit to the data (χ2 (14) = 95.772, *p*<.001), correctly discriminated 63.6% of cases and Nagelkerke R^2^ indicated a 13% variation of stress explained by the model. Table 4 presents results of the multivariable logistic regression.

**Table 4.**
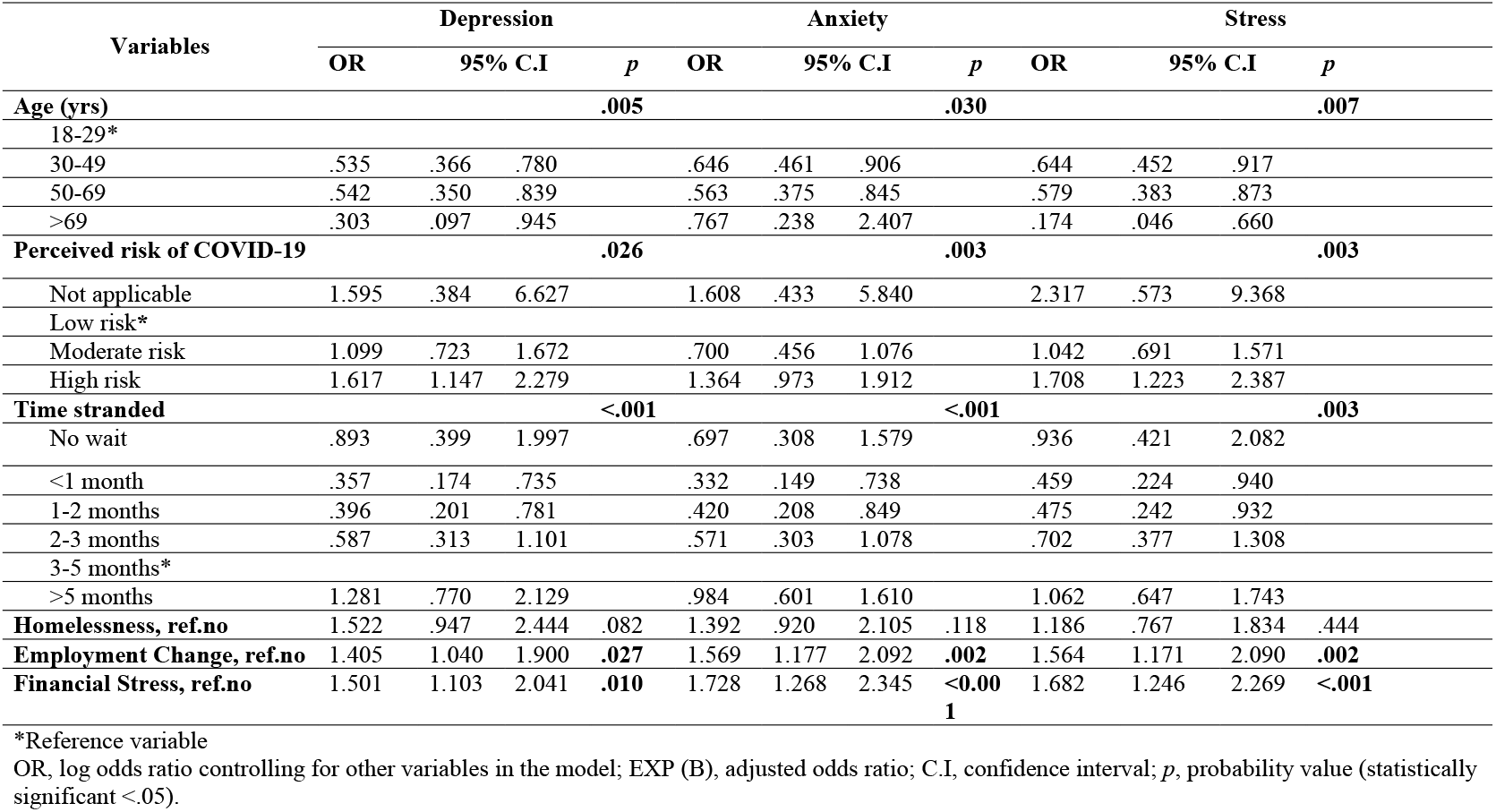
Predictors of moderate to extremely severe depression, anxiety, and stress in participants stranded abroad during COVID-19 (n=956)

Being 30 years or older and stranded for 2 months or less was associated with decreased odds of moderate to extremely severe depression, anxiety, and stress.

## DISCUSSION

This study evaluated the psychological and financial distress of individuals stranded abroad during the COVID-19 pandemic and highlights the importance of providing additional support to this vulnerable group in future public health events. Amongst our respondents we found that over half had been stranded for longer than five months, with the majority having more than one flight cancellation or change. Our results confirm sentiments shared on social media by people stranded abroad, that reflect experiences of having no financial support, depression, homelessness and a general feeling of abandonment by their governments (7-9). Given the continued flight changes and delays (incurring additional costs) in the population of individuals stranded abroad, along with changes to employment, it is perhaps not surprising that we documented a high level of financial distress (64.2%), employment changes (45%), and experiences of homelessness (12%). Our findings align with results from non-travel related COVID studies which have indicated an increase in financial distress (16), increases in experiences of homelessness (26) and growing employment changes (27) during the pandemic. Comparatively, we reported much higher findings compared to a survey of the general population conducted within the first 6 months of the pandemic, finding that 30% of Australians were financially stressed because of the pandemic (28). This difference could be explained by those stranded abroad having different elements of uncertainty (additional flight costs, additional rent, and expenses due to the length of time stranded, and uncertain employment) compared to the general population.

Based on our findings, we recommend policymakers prioritise increasing the availability of financial assistance in the form of government grants or loans for living and flight costs incurred due to being stranded abroad, or alternatively providing the option to access social support while abroad if an individual would have been eligible had they not been abroad. Furthermore, considering the proportion of people who reported experiencing homelessness, the cases of sexual assault within homeless shelters, and previous research indicating a lack of emergency accommodation options for citizens abroad during COVID-19 (6), it is recommended that policymakers provide a solution to this issue, whether it be through financial assistance or an emergency accommodation program similar to those that the French and Spanish governments introduced (6).

At this stage in the pandemic, it is almost indisputable that COVID-19 has had a psychological impact on populations around the world, whether it be healthcare workers, people in lockdown or quarantine, or specific countries or communities, the stressors were all encompassing (10-13, 29, 30). Our findings reflect much higher incidence of moderate to extremely severe depression (64.4%), anxiety (41.7%), and stress (58.1%), compared to domestic populations around the world (24). Previous research show lower DAS severity scores (24), especially when comparing our results to studies exploring psychological impacts of COVID-19 on healthcare workers (31, 32) and domestic students (33, 34). One study observing the psychological correlates of COVID-19 on the general population in Austria, reported drastically lower scores of depression (21.6%), anxiety (28.6%) and stress (28%), and found that ‘frequent contact with family or friends’ was shown to be a protective factor (24).

However, research on international students during COVID-19, have reflected high DASS severity, similar to our study findings (19, 35). Possible reasons for these differences could be the parallels between international student experiences and those stranded, both living abroad and arguably away from their immediate social support network (family). A range of factors contributed to the psychological wellbeing of individuals in this study. Having financial stress, any employment change, being aged <50yrs, having a high perceived risk of contracting COVID-19 and being stranded for >5months were all associated with predicting moderate to extremely severe depression, anxiety, and stress. These results are not surprising considering the literature shows that many stressors, like financial distress, fear of COVID-19 infection, loneliness, inadequate information, and employment issues have all presented as predictors of poor mental health and in the case of financial distress, can go further than predicting depression to suicidal thoughts and behaviours (30, 36). Unique to this study, however, is the finding that the longer an individual is stranded abroad the more likely they are to present with moderate to severe DAS symptoms.

Interestingly, of the age categories in this investigation, participants >69yrs had lower scores of depression, anxiety, and stress, inconsistent with research conducted in Spain and Canada suggesting increased DASS scores in elders (37, 38). This, as was noted by the authors of the Sightlines Project (39), may be due to older people being more financial secure than other age groups. It may also reflect that older people were less likely to have insecurity associated with employment or have younger dependent family members to provide direct care to. This may have provided more opportunity for flexibility in their travel plans.

These results reflecting high severity of DASS in those stranded abroad, provides both current and future direction for policymakers. We recommend policymakers provide adequate mental health interventions be available to those stranded abroad, either online or face-to-face where possible through a local consulate.

It is hard to deny that people have been deprived of the ability to return to their country of citizenship or permanent residence, as shown by the 63.3% of our respondents being stranded abroad for longer than 5 months. Addressing public health threats from a health security perspective has already increased fears that it legitimises government actions, potentially undermining personal sovereignty, and impeding human rights (40). A commonly cited human rights treaty in response to imposed restrictions is Article 12 (41) of the International Covenant on Civil and Political Rights (ICCPR), a treaty with 74 signatories and 173 parities which states that “No one shall be arbitrarily deprived of the right to enter his own country” (41).

The most frequently mentioned human rights breach was the Australian government imposing not only a complete ban on incoming flights from India, but potential criminal penalties of up to five years imprisonment, and/or fines up to $66000 (AUD) (42). Mostly in reference to breaching Article 12 (41) of the International Covenant on Civil and Political Rights (ICCPR), this ban, implemented under the *Biosecurity Act 2015(43)*, between April and May 2021, has been labelled a ‘racist rights breach’ (44), with the Australian Human Rights Commission approaching the federal government directly with their concerns (42). Due to impacts of ongoing border closures and individuals struggling to return home, as further highlighted in this study, Australian citizens stranded abroad submitted a human rights complaint to the United Nations Human Rights Committee under the ICCPR, with the UN Human Rights Committee already successfully requesting to the Australian Government the prompt repatriation of two Australians in April 2021 (45).

Due to the results of our study alongside the probable human rights breaches, we recommend policymakers seriously reconsider current and future restriction of movement of citizens and permanent residents returning home who are at risk of financial distress and severe DASS the longer that they are stranded for. With international borders reopening around the world, some being restricted for nearly 20 months, it is critical to not only look at the impact of travel restrictions from the perspective of reducing infectious disease importations, but also from the perspective of those stranded abroad, who were arguably one of the most impacted by them. In doing so, policymakers can determine where further support is needed in future emergency situations resulting in people stranded abroad.

## Limitations

This work is not without limitations. Firstly, like other cross sectional survey studies, it lacks a longitudinal follow up on participants. Secondly, the self-report questionnaire for psychological symptoms raises possible selection bias and subjectivity, however our sample size being large, and the addition of an optional ‘do not recall’ response should mitigate certain bias. Thirdly, we were unable to capture the specific immigration status of those returning, i.e., whether they were citizens, permanent residents, or short-term visa holders. Finally, we did not examine pre-existing mental health conditions which could prove to be a confounding factor, and there was an over representation of women, those returning to the western pacific region and participants with an academic background, possible due to convenience sampling issues and survey distribution from Australia, which may not fully represent the population of people who were stranded abroad. Notwithstanding the previously mentioned limitations, findings from this study provide insights not previously reported into the psychological and financial impacts and support needed for individuals stranded abroad due to COVID-19 travel restrictions. These insights will be valuable for policy makers as they design and deliver support programs in response and prepare for future events.

## Conclusion

This research suggests that being stranded abroad during COVID-19 may lead to not only an increase in financial stress, but quite severe depression, anxiety, and stress. Our findings show that being young, stranded abroad for longer, having a high perception of infection risk, experiencing employment changes and financial stress are all associated with increased severity of depression, anxiety, and stress. Participants reported that lack of mental and social health support while stranded. This indicates that there are gaps in services available for this vulnerable population or lack of communication as to how to access them; both issues that need to be resolved.

## Data Availability

All data produced in the present study are available upon reasonable request to the authors

## ABBREVIATIONS

AFR: African region
AMR: Region of the Americas
COVID-19: Novel coronavirus disease 2019
DAS: Depression, anxiety, and stress
DASS-21: The 21-item depression, anxiety, and stress scale
EMR: Eastern Mediterranean region
EUR: European region
SEAR: South-East Asian region
WPR: Western Pacific region

## DECLARATIONS

### Ethics approval and consent to participate

Ethical approval for this study was granted by the UNSW Human Research Ethics Committee (#210418). All participants were anonymous and indicated their consent to participate.

## Supplementary information

**Supplementary file 1**.

Associations between participant factors and levels of depression, anxiety, and stress

